# Depressive symptoms and Prevalent Cardiovascular Disease: A Cross-Sectional Analysis in Port-au-Prince, Haiti

**DOI:** 10.64898/2026.07.15.26358073

**Authors:** Daniella Myriam Pierre, Rehana Rasul, Reichling St. Sauveur, Kelly Celestin, Vanessa Rouzier, Erline Hilaire, Marie Marcelle Deschamps, Jean William Pape, Lily D. Yan, Anju Ogyu, Catherine Bennett, Margaret L. McNairy, Rodney Sufra, Denis Nash

## Abstract

**Background:** Cardiovascular disease (CVD) is the leading cause of mortality in low- and middle-income countries (LMICs). In Haiti, depression remains an underexplored CVD risk factor. We assessed the association between depressive symptoms (DS) and prevalent CVD in urban Haiti and examined sex differences.

**Methods:** We conducted a cross-sectional analysis of enrollment data from the Haiti Cardiovascular Disease Cohort (adults ≥18 years; March 2019–August 2021). DS were measured using the Patient Health Questionnaire-9 (PHQ-9) and categorized as none–mild (<10) versus moderate–severe (≥10). Prevalent CVD (angina, myocardial infarction, transient ischemic attack or stroke, heart failure) was adjudicated using epidemiologic definitions aligned with international cohorts. We estimated prevalence ratios (PRs) using generalized estimating equation Poisson models with a log link, adjusting for age, sex, education, income, food insecurity, smoking, alcohol use, physical activity, stress, and BMI. Effect modification by sex was assessed on multiplicative and additive scales.

**Results:** Among 2,995 participants (mean age 41.9 years; 58.0% female), 16.2% (95% CI: 14.8–17.5; n=484) had moderate–severe DS. Prevalence was higher in females (22.0%, 95% CI: 19.8–23.6) than males (8.5%, 95% CI: 7.0–10.1). The prevalence of CVD was higher among participants with moderate–severe DS compared with those with none–mild DS, with similar patterns observed in both sexes (males: 21.5% vs 10.6%; females: 23.3% vs 15.3%). Moderate–severe DS were associated with higher CVD prevalence compared with none–mild DS (adjusted PR [aPR]=1.36; 95% CI: 1.08–1.71). In sex-stratified models, aPRs were 1.38 (95% CI: 1.06–1.78) for females and 1.25 (95% CI: 0.75–1.99) for males. Evidence for interaction by sex on the additive scale was limited (RERI=0.07, 95% CI: −0.74 to 0.88).

**Conclusion:** Moderate-to-severe DS were independently associated with a higher prevalence of CVD in urban Haiti. Associations were consistently stronger among women, although evidence for effect modification by sex was limited. Integrating depression screening and management into CVD prevention efforts may help address the growing burden of both conditions in resource-limited settings. Prospective studies are warranted to better understand the underlying mechanisms and causal pathways.

## Introduction

Cardiovascular diseases (CVD) are the leading cause of morbidity and mortality globally.^1^ In low- and middle-income countries (LMIC) like Haiti, CVD accounts for an estimated 36% of adult deaths, surpassing infectious diseases as the most common cause of death.^1,2^ Beyond traditional risk factors such as hypertension, diabetes, and obesity, CVD prevalence in impoverished settings is strongly influenced by social determinants of health (SDoH), including economic inequities, food insecurity, and poverty-related stress. These factors may further exacerbate CVD risk and worsen mental health disorders.^1,3^

Depression is associated with an increased risk of CVD through a combination of physiological, behavioral and social pathways.^4^ Physiologically, depression can activate the hypothalamic-pituitary-adrenal axis, contribute to nervous system autonomic dysregulation, and increase systemic inflammation, all of which have been linked to the development and progression of cardiovascular conditions.^5^ Behaviorally, depression may reduce adherence to cardiovascular medications and hinder engagement in health-promoting behaviors such as physical activity, balanced nutrition, and smoking cessation. Socially, individuals with depression often experience lower levels of social support, increased isolation, and reduced access to care, further compounding CVD risk.^2,3,6^ In Haiti, these effects may be intensified by persistent social and structural challenges, including food insecurity, limited access to mental health services, and widespread poverty^3,6^

Women in LMICs face higher prevalence of both depression^7^ and CVD compared to men.^4,8^ Depression prevalence rise significantly in women after mid-puberty, potentially due to hormonal and psychosocial factors.^4,9^ However, sex differences in the effect of depression on CVD remain inconclusive.^10–13^ This gap emphasizes the need to explore sex-specific dynamics in CVD risk within culturally unique contexts.

Although studies in high-income countries support the depression - CVD relationship,^14,15^ data from LMICs including Haiti remain limited.^3,4^ In Haiti, chronic humanitarian crises may exacerbate the burden of both conditions, yet few studies have investigated their interplay.

To address this gap, this study investigates the relationship between depressive symptoms (DS) and CVD in a community-based urban cohort study of adults living in Port-au-Prince, Haiti.

Specifically, we aim to estimate the association between DS and CVD prevalence and examine whether sex modifies this relationship. We hypothesize that a higher level of DS is associated with greater CVD prevalence, with a stronger effect in females than in males.

## Methods

### Ethics Statement

Before enrollment, all participants provided written informed consent. The study received input from community stakeholders and GHESKIO’s Community Advisory Board prior to its initiation. The institutional review boards at Weill Cornell Medicine and GHESKIO in Haiti approved all study protocols and procedures. Additional information regarding the ethical, cultural, and scientific considerations specific to inclusivity in global research is included in the Supporting Information (S1 Checklist).

### Study design and setting

The Haiti Cardiovascular Disease Cohort Study is a longitudinal community-based urban cohort which seeks to investigate the prevalence and incidence of CVD and risk factors in Port-au-Prince, Haiti.^1^ The study is conducted by the Groupe Haïtien d’Etude du Sarcome de Kaposi et des Infections Opportunistes (GHESKIO), a medical non-profit organization established in 1982 to provide clinical care and actionable research on infectious and non-communicable diseases.^1^ Participants were selected using a multistage random sampling design in Port-au-Prince, guided by GPS waypoints distributed across census blocks. The number of waypoints per block was proportional to the estimated population size, as determined by data from the Institut Haïtien de Statistique et d’Informatique.^16^ Eligibility required adults (aged 18 or older) whose primary residence was in Port-au-Prince without any cognitive impairments or severe medical conditions.^1^ This cross-sectional analysis of the Haiti Cardiovascular Disease Cohort Study used enrollment data from all participants recruited between March 2019 and August 2021. The study population comprised 3,005 adults (Fig S1). The final sample comprised 2,995 after excluding participants who missed the PHQ-9 assessment (n=10). The study followed the Strengthening the Reporting of Observational Studies in Epidemiology (STROBE) statement^17^ for cross sectional studies.

### Characteristics of Depressive symptoms (DS)

DS level was measured using a Creole translated version of the Patient Health Questionnaire 9 (PHQ-9), a validated instrument commonly used to measure DS and is widely used in clinical and research contexts with high validity and reliability.^18,19^ The PHQ-9 consists of 9 items related to DS, with response options ranging from 0 (not at all) to 3 (nearly every day). Responses were summed to give a total symptom score ranging from 0 to 27.^3,19^ DS severity was categorized as: “none to mild” (score < 10) and “moderate to severe” (score ≥10).^20^ Internal consistency of the PHQ-9 tool was assessed using Cronbach’s alpha, which demonstrated good internal consistency in this sample (α = 0.78). This reliability estimate is comparable to values reported in U.S. populations, where Cronbach’s alpha typically ranges from approximately 0.79‒0.93 across diverse clinical and community settings.^21–24^

### Dependent variable: CVD

The dependent variable was prevalent CVD (Yes/No), which was defined as a composite of any of the following events: heart failure, stroke, transient ischemic attack, myocardial infarction, and angina using epidemiological criteria from the American Heart Association, the WHO and the European Society of Cardiology, and were adjudicated by a three-physician adjudication committee and classified as definite, possible and probable based on a review of patient-reported symptoms, medical history, physical exam findings, imaging data, and laboratory results, following similar methods used in the REasons for Geographic and Racial Differences in Stroke (REGARDS) and Coronary Artery Risk Development in Young Adults Study (CARDIA) studies.^2,25,26^ Only the definite and probable events were included in the present analysis. Detailed CVD definitions, adjudication procedures, and classification criteria are provided in Supporting Information (Appendix S1 Table 1) and prevalence of each CVD (Appendix S1 Table 2) and were previously published for the Haiti Cardiovascular Disease Cohort.^2^

**Table 1.**
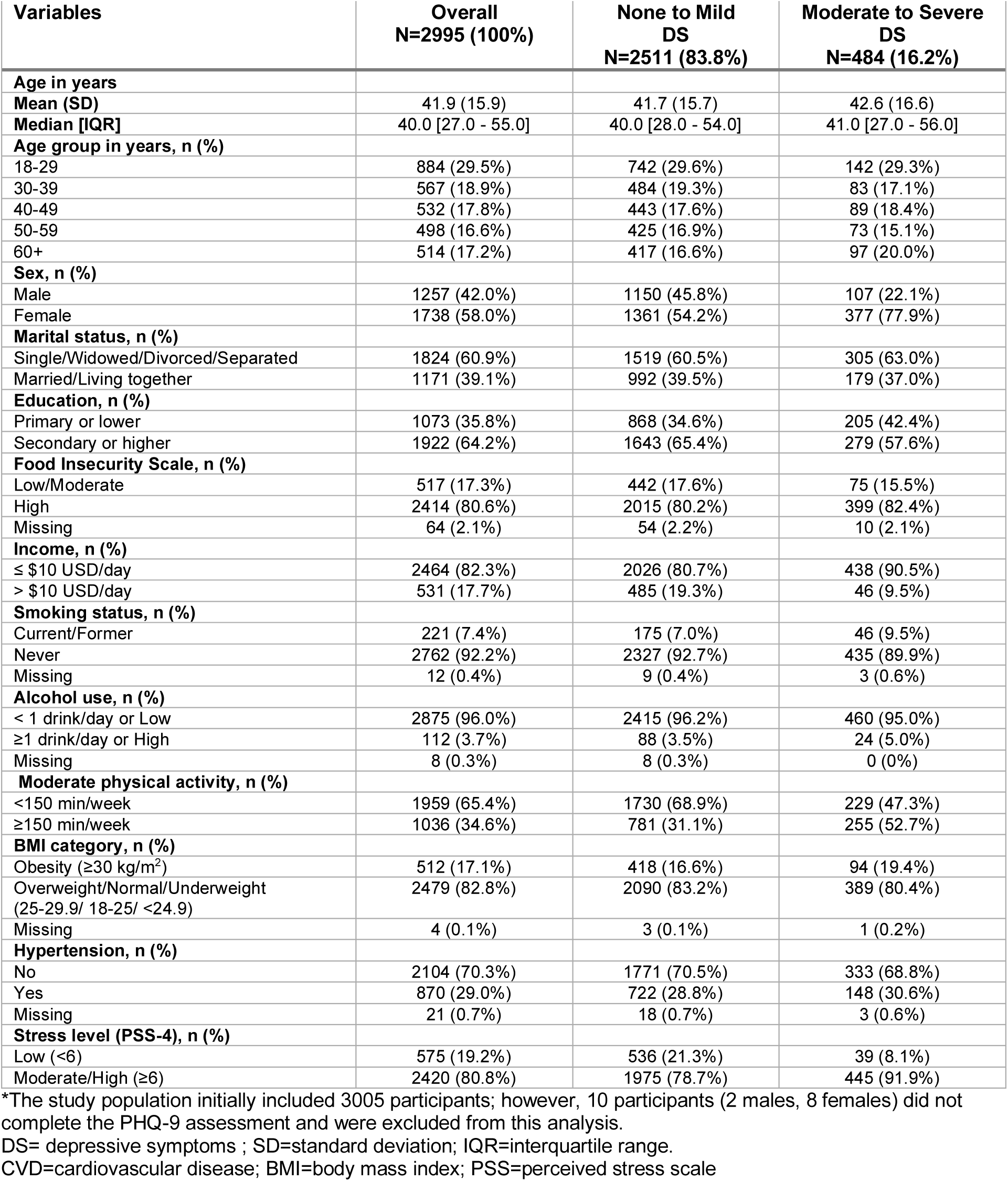
Participant characteristics by DS status (n=2995)* in the HCVD Cohort, March 2019-August 2021, Port-au-Prince, Haiti.

| Variables | Overall<br>N=2995 (100%) | None to Mild<br>DS<br>N=2511 (83.8%) | Moderate to Severe<br>DS<br>N=484 (16.2%) |
| --- | --- | --- | --- |
| **Age in years** |  |  |  |
| Mean (SD) | 41.9 (15.9) | 41.7 (15.7) | 42.6 (16.6) |
| Median [IQR] | 40.0 [27.0 - 55.0] | 40.0 [28.0 - 54.0] | 41.0 [27.0 - 56.0] |
| **Age group in years, n (%)** |  |  |  |
| 18-29 | 884 (29.5%) | 742 (29.6%) | 142 (29.3%) |
| 30-39 | 567 (18.9%) | 484 (19.3%) | 83 (17.1%) |
| 40-49 | 532 (17.8%) | 443 (17.6%) | 89 (18.4%) |
| 50-59 | 498 (16.6%) | 425 (16.9%) | 73 (15.1%) |
| 60+ | 514 (17.2%) | 417 (16.6%) | 97 (20.0%) |
| **Sex, n (%)** |  |  |  |
| Male | 1257 (42.0%) | 1150 (45.8%) | 107 (22.1%) |
| Female | 1738 (58.0%) | 1361 (54.2%) | 377 (77.9%) |
| **Marital status, n (%)** |  |  |  |
| Single/Widowed/Divorced/Separated | 1824 (60.9%) | 1519 (60.5%) | 305 (63.0%) |
| Married/Living together | 1171 (39.1%) | 992 (39.5%) | 179 (37.0%) |
| **Education, n (%)** |  |  |  |
| Primary or lower | 1073 (35.8%) | 868 (34.6%) | 205 (42.4%) |
| Secondary or higher | 1922 (64.2%) | 1643 (65.4%) | 279 (57.6%) |
| **Food Insecurity Scale, n (%)** |  |  |  |
| Low/Moderate | 517 (17.3%) | 442 (17.6%) | 75 (15.5%) |
| High | 2414 (80.6%) | 2015 (80.2%) | 399 (82.4%) |
| Missing | 64 (2.1%) | 54 (2.2%) | 10 (2.1%) |
| **Income, n (%)** |  |  |  |
| ≤ \$10 USD/day | 2464 (82.3%) | 2026 (80.7%) | 438 (90.5%) |
| > \$10 USD/day | 531 (17.7%) | 485 (19.3%) | 46 (9.5%) |
| **Smoking status, n (%)** |  |  |  |
| Current/Former | 221 (7.4%) | 175 (7.0%) | 46 (9.5%) |
| Never | 2762 (92.2%) | 2327 (92.7%) | 435 (89.9%) |
| Missing | 12 (0.4%) | 9 (0.4%) | 3 (0.6%) |
| **Alcohol use, n (%)** |  |  |  |
| < 1 drink/day or Low | 2875 (96.0%) | 2415 (96.2%) | 460 (95.0%) |
| ≥1 drink/day or High | 112 (3.7%) | 88 (3.5%) | 24 (5.0%) |
| Missing | 8 (0.3%) | 8 (0.3%) | 0 (0%) |
| **Moderate physical activity, n (%)** |  |  |  |
| <150 min/week | 1959 (65.4%) | 1730 (68.9%) | 229 (47.3%) |
| ≥150 min/week | 1036 (34.6%) | 781 (31.1%) | 255 (52.7%) |
| **BMI category, n (%)** |  |  |  |
| Obesity (≥30 kg/m<sup>2</sup>) | 512 (17.1%) | 418 (16.6%) | 94 (19.4%) |
| Overweight/Normal/Underweight (25-29.9/ 18-25/ <24.9) | 2479 (82.8%) | 2090 (83.2%) | 389 (80.4%) |
| Missing | 4 (0.1%) | 3 (0.1%) | 1 (0.2%) |
| **Hypertension, n (%)** |  |  |  |
| No | 2104 (70.3%) | 1771 (70.5%) | 333 (68.8%) |
| Yes | 870 (29.0%) | 722 (28.8%) | 148 (30.6%) |
| Missing | 21 (0.7%) | 18 (0.7%) | 3 (0.6%) |
| **Stress level (PSS-4), n (%)** |  |  |  |
| Low (<6) | 575 (19.2%) | 536 (21.3%) | 39 (8.1%) |
| Moderate/High (≥6) | 2420 (80.8%) | 1975 (78.7%) | 445 (91.9%) |

\*The study population initially included 3005 participants; however, 10 participants (2 males, 8 females) did not complete the PHQ-9 assessment and were excluded from this analysis.
DS= depressive symptoms ; SD=standard deviation; IQR=interquartile range.
CVD=cardiovascular disease; BMI=body mass index; PSS=perceived stress scale

**Table 2.**
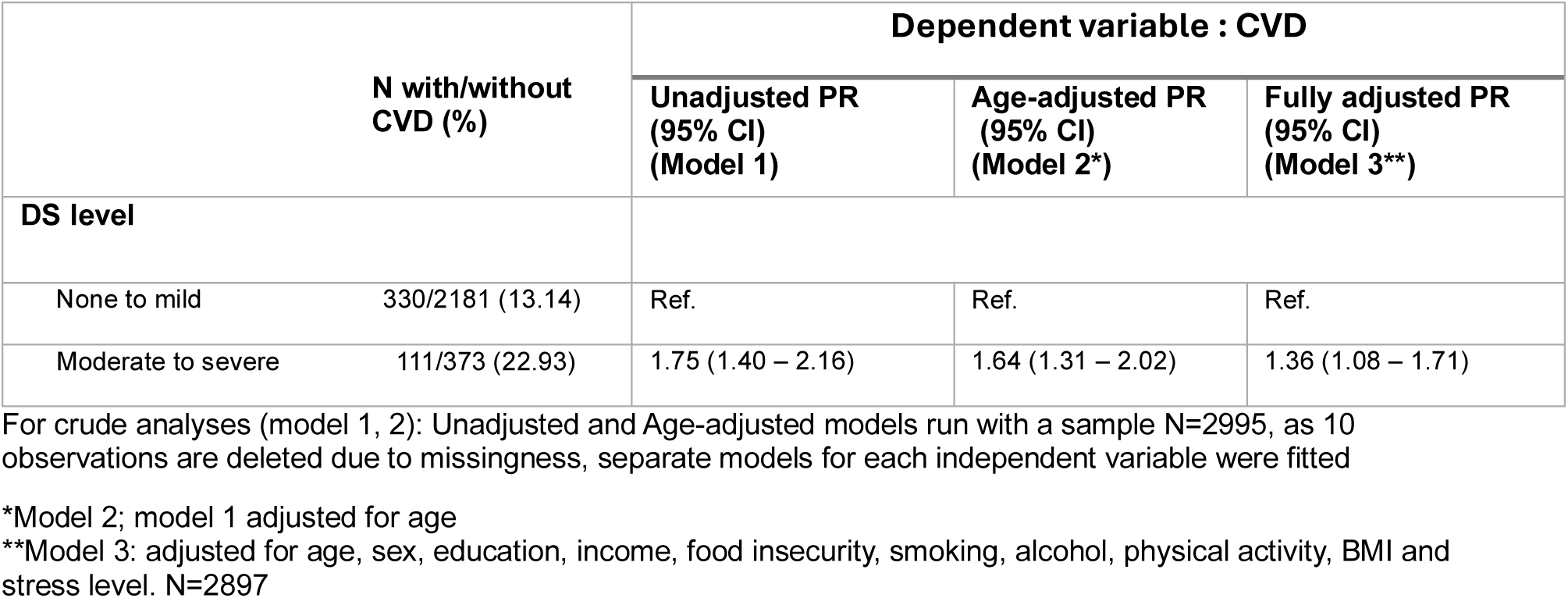
Unadjusted, Age-Adjusted and Fully Adjusted PRs for Depressive Symptoms (DS) -CVD Association in HCVD Cohort, 2019-2021, Port-au-Prince, Haiti.

### Confounders and other measures

Potential confounders of the relationship between DS and CVD were identified using a directed acyclic graph (DAG)^27^ depicting the hypothesized relationship between DS and CVD (Fig. S2). The causal structure of the DAG was informed by prior epidemiologic evidence demonstrating that DS influence CVD risk through behavioral and metabolic pathways. Traditional cardiovascular risk factors such as hypertension, diabetes, and dyslipidemia may lie on the causal pathway between DS and CVD; therefore, these variables were not included in the primary adjustment set to avoid potential overadjustment.^28–30^ Physical activity and BMI were included in the primary adjustment set based on the hypothesized DAG; however, because these variables may also lie on the causal pathway between depressive symptoms and cardiovascular disease, adjustment for them may estimate an association closer to a controlled direct effect rather than the total association. Therefore, we conducted a sensitivity analysis excluding physical activity and BMI from the adjusted model. These confounders included age in years, socioeconomic status : education level (primary or lower, secondary or higher), income (≤ $10 USD/day, >$10 USD/day for the household), household food insecurity (low or moderate, high) based on the six-item questionnaire for the Household Food Security Scale,^31,32^ WHO STEPwise Approach to Non-communicable Disease Risk Factor Surveillance instrument was used to measure lifestyle factors included smoking status (current or former, never) assessed through questions related to history or current use of tobacco products, alcohol use within the last 12 months (<1 drink/day, ≥1 drink/day) and physical activity (< 150 minutes per week of moderate to vigorous exercises to entertain or for fitness, ≥ 150 minutes per week of moderate to vigorous exercises to entertain or for fitness), body mass index (BMI) category (obesity (BMI) ≥ 30 kg/m^2^ or overweight/normal weight/underweight (25-29.9/ 18.5-24.9/ <18.5) and stress measured using the Cohen’s Perceived Stress Scale 4/PSS-4 (low (PSS-4 < 6), moderate (≥ 6 PSS-4 ≤ 10) or high (PSS-4 ≥ 11).^2,33^

Marital status (single or widowed or divorced or separated, married or living together) was assessed as a sociodemographic characteristic. Hypertension (yes/no) defined based on the average of the last two of three measurements of systolic (SBP ≥ 140 mmHg) and diastolic (DBP ≥ 90 mmHg) blood pressure by a healthcare provider during clinic visits or self-reported of taking antihypertensive medications^34^, was considered a potential mediator variable in the causal pathway between DS and CVD consistent with prior studies demonstrating associations between DS or depression and incident hypertension and the contribution of hypertension to cardiovascular risk, although the extent of mediation remains uncertain.^28,35,36^

### Effect modifier

Given evidence in the literature on sex differences suggesting that females may be more susceptible to the cardiovascular effects of depression than males,^10,11^ sex was examined as an effect modifier of the relationship between depression and CVD on the additive and multiplicative scales.

### Statistical methods

R software (R 4.4.1)^37^ was used for statistical analysis. Participant characteristics were described by DS level and sex. DS were categorized as none to mild versus moderate to severe for the primary analysis and additionally categorized into three levels (none: score ≤ 4, mild: score 5 - 9, and moderate to severe ≥ 10) to assess the robustness of the main findings. The association between DS and CVD was measured using unadjusted, age-adjusted and fully adjusted multivariable generalized estimating equations (GEE) with a Poisson distribution and a log link to account for clustering of participants by household. Fully adjusted models (multivariable) included the following confounders based on directed acyclic graphs (DAG) used to identify the covariates required to adjust for (Figure S2A): age, sex, income, education, food insecurity, smoking, alcohol, physical activity, BMI, and stress. Hypertension likely mediates the relationship between depression and CVD, thus was not adjusted for in our models. Multicollinearity was evaluated using the variance inflation factor (VIF), with VIF>5 indicating multicollinearity.

Effect measure modification (EMM) between sex and DS on CVD was estimated on the additive and multiplicative scales (Supporting information, Statistical appendix), and stratified analyses were performed to assess differences between strata as sensitivity analysis.^38–40^ Positive additive interaction was defined as Relative Excess Risk due to Interaction (RERI)> 0. The 95% CI for RERI was generated by using the spreadsheet tool reported by Knol and VanderWeele.^41^ Multiplicative EMM, which is present when the combined effect of two exposures exceeds (or falls short of) the product of their individual effects, was estimated by including an interaction term (DS*sex) to the adjusted multivariable Poisson regression model. (Supporting information, Statistical appendix)

### Missing data & sensitivity analysis

Missing data were assessed for all variables included in the analytic model. DS were missing for 10 participants (0.3%; 8 males and 2 females), while no missing data were observed for the dependent variable CVD. Missingness in covariates resulted in the exclusion of 98 participants (3.3% of the analytic sample) in the fully adjusted models. Given the low proportion of missing data, the primary analysis used complete-case data. To evaluate the potential impact of missing data on study findings, under the assumption that data were missing at random, a sensitivity analysis using multiple imputation by chained equations was performed with the *mice* package in R. Twenty imputed datasets were generated, including the dependent variable, exposure, and all covariates in the analytic model. Models were fitted within each imputed dataset, and estimates were combined using Rubin’s rules.^42^

## Results

### Participants characteristics

The study included 2,995 participants, of whom 484 (16.2%) were classified as having moderate to severe DS. DS were missing for 10 participants (0.3%; 8 males and 2 females), while no missing data were observed for the CVD dependent variable. The mean age was 42.0 (SD=15.9) years old. Females comprised 78.0% of the moderate to severe DS group, (Table 1, Fig1). Across age groups, the prevalence of moderate to severe DS remained consistently higher among females than males. (Table S1, Fig.S3).

Participants with moderate to severe DS were more likely to have primary or lower education levels (42.4%), high food insecurity (82.4%, and household income of $10 USD or less per day (90.5%). Regarding lifestyle factors, moderate physical activity was more common among individuals with moderate to severe DS (53.0%), while obesity and hypertension prevalence were slightly higher among those with moderate to severe DS. Finally, moderate to high stress levels were more prevalent among participants with moderate to severe DS (91.9%). (Table 1, Table S1)

### Multivariable adjusted association between DS and prevalent CVD

The fully adjusted models excluded 98 participants (3.3% of the analytic sample) due to missing covariate data, resulting in a final analytic sample of 2,897 participants. After adjusting for confounders, participants with moderate to severe DS had a higher prevalence ratio (PR) of CVD compared to those with none to mild DS (aPR = 1.36 (95% CI: 1.08 –1.71). (Table 2)

Results for effect measure modification (EMM) by sex are presented in Table 3 and 4. Moderate to severe DS was associated with a higher prevalence of CVD in both sexes, with an aPR of 1.25 (95% CI: 0.75 – 1.99) for males and 1.38 (95% CI: 1.06 –1.79) for females. No evidence of effect measure modification by sex was found on either the multiplicative scale (interaction p-value > 0.9) or the additive scale (RERI = 0.07, 95% CI: −0.74 to 0.88). (Table 4)

**Table 3.** Association between Depressive Symptoms (DS) severity (three categories) and CVD by Adjustment Level and Sex in HCVD Cohort, Haiti.

| Models | Unadjusted Model | Age-adjusted Model | Adjusted Model 2* |
| --- | --- | --- | --- |
|  | PR (95%CI) | PR (95%CI) | PR (95%CI) |
| **DS level** |  |  |  |
| **Overall** |  |  |  |
| None | Ref. | Ref. | Ref. |
| Mild | 1.27 (1.02 – 1.57) | 1.26 (1.02 – 1.57) | 1.14 (0.91 – 1.44) |
| Moderate to severe | 1.92 (1.51 – 2.42) | 1.80 (1.42 – 2.27) | 1.45 (1.12 – 1.87) |
| **Among Males†** |  |  |  |
| None |  |  | Ref. |
| Mild |  |  | 1.04 (0.69 – 1.54) |
| Moderate to severe |  |  | 1.37 (0.80 – 2.24) |
| **Among Females†** |  |  |  |
| None |  |  | Ref. |
| Mild |  |  | 1.19 (0.89 – 1.58) |
| Moderate to severe |  |  | 1.49 (1.09 – 2.02) |
| **Measure of EMM on multiplicative scale\*\* & Interaction term p-value:**<br>Mild DS*Female aPR (95% CI) = 1.06 (0.71 – 1.56); P = 0.7<br>Moderate to severe DS*Female aPR (95% CI) = 1.36 (0.81 – 2.19); p-value = 0.7 |  |  |  |

PR= prevalence ratio, CI=confidence interval
\*Model 2 from a generalized estimating equation Poisson regression distribution fully adjusted for age, sex, education, income, food insecurity, alcohol, smoking, physical activity and obesity
\*\*Model 3 from a generalized estimating equation Poisson regression distribution with DS\*sex interaction and adjusted for age, sex, education, income, food insecurity, alcohol, smoking, physical activity and obesity Interaction term DS\*sex p-value=0.7 (for both subcategory Mild DS\*Female, and Moderate to severe DS\*Female)

**Table 4.**
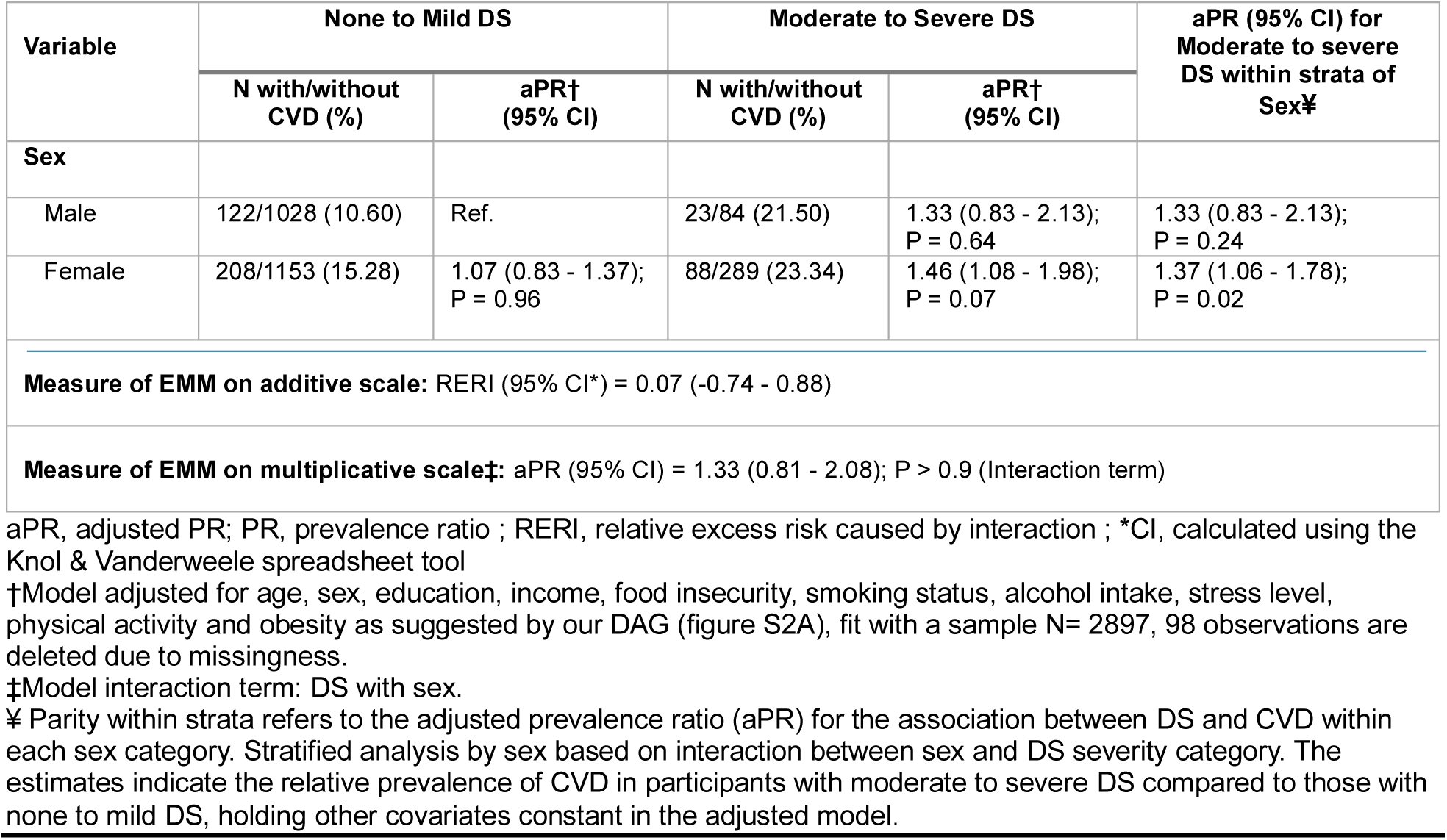
Effect Modification by Sex of the Depressive Symptoms (DS) -CVD Association in HCVD Cohort (N=2897), 2019-2021, Port-au-Prince, Haiti.

| Variable | None to Mild DS |  | Moderate to Severe DS |  | aPR (95% CI) for Moderate to severe DS within strata of Sex‡ |
| --- | --- | --- | --- | --- | --- |
|  | N with/without CVD (%) | aPR† (95% CI) | N with/without CVD (%) | aPR† (95% CI) |  |
| **Sex** |  |  |  |  |  |
| Male | 122/1028 (10.60) | Ref. | 23/84 (21.50) | 1.33 (0.83 - 2.13);<br>P = 0.64 | 1.33 (0.83 - 2.13);<br>P = 0.24 |
| Female | 208/1153 (15.28) | 1.07 (0.83 - 1.37);<br>P = 0.96 | 88/289 (23.34) | 1.46 (1.08 - 1.98);<br>P = 0.07 | 1.37 (1.06 - 1.78);<br>P = 0.02 |
| **Measure of EMM on additive scale:** RERI (95% CI*) = 0.07 (-0.74 - 0.88) |  |  |  |  |  |
| **Measure of EMM on multiplicative scale‡:** aPR (95% CI) = 1.33 (0.81 - 2.08); P > 0.9 (Interaction term) |  |  |  |  |  |

aPR, adjusted PR; PR, prevalence ratio ; RERI, relative excess risk caused by interaction ; \*CI, calculated using the Knol & Vanderweele spreadsheet tool
†Model adjusted for age, sex, education, income, food insecurity, smoking status, alcohol intake, stress level, physical activity and obesity as suggested by our DAG (figure S2A), fit with a sample N= 2897, 98 observations are deleted due to missingness.
‡Model interaction term: DS with sex.
‡ Parity within strata refers to the adjusted prevalence ratio (aPR) for the association between DS and CVD within each sex category. Stratified analysis by sex based on interaction between sex and DS severity category. The estimates indicate the relative prevalence of CVD in participants with moderate to severe DS compared to those with none to mild DS, holding other covariates constant in the adjusted model.

The analysis using three-level DS categories confirmed that moderate to severe DS was consistently associated with higher CVD prevalence, particularly among females (PR = 1.50; 95% CI: 1.11–2.02). And there was no evidence of association for mild DS and CVD in any model or sex-strata (Table 3). For the sensitivity analysis using multiple imputation, the pooled prevalence ratios from the imputed datasets were almost identical to estimates from the complete case analysis (PR 1.37, 95% CI 1.09–1.71, p=0.006 vs. PR 1.36, 95% CI 1.08–1.72, p=0.008). Lastly the sensitivity analysis excluding physical activity and BMI from the adjusted model yielded slightly stronger associations between moderate-to-severe DS and prevalent CVD (aPR = 1.48; 95% CI: 1.18–1.85), while the overall interpretation of the findings remained unchanged (Supporting information, Table S2).

## Discussion

In this community-based urban cohort of 2,995 adults living in Haiti, DS prevalence was 16.2%, and it was higher among individuals facing greater socio-economic disadvantage and stress. We also found that moderate to severe DS was significantly associated with CVD, even after adjusting for sociodemographic, behavioral, and clinical factors. Although we observed stronger associations among women, there was no statistically significant effect modification by sex.

Depression is a growing public health concern in Haiti, with prevalence patterns similar to or higher than those reported in other LMICs. Evidence from both Haiti and other LMICs consistently links elevated depression rates to intersecting stressors, including food insecurity, violence, displacement, and weak health systems.^2,3,6^ These challenges are particularly chronic in Haiti, where ongoing political unrest and escalating gang violence have likely intensified the mental health crisis in recent years.^3^ Access to mental health care remains limited, and many individuals are unaware that depression and related conditions are treatable which further compound the burden of untreated mental illness.

Our findings reinforce a growing body of evidence that identifies depression as an independent risk factor for CVD beyond traditional CVD risk factors.^43^ Previous studies, largely based in high-income countries and predominantly white populations, have linked depression to increased CVD risk through behavioral and biological mechanisms, such as systemic inflammation, hypothalamic-pituitary-adrenal axis dysregulation, and comorbid conditions like hypertension and diabetes. Sensitivity analyses excluding physical activity and obesity yielded stronger associations, suggesting that these factors may partially mediate or attenuate the relationship between DS and CVD. Although limited research has explored these associations in Black populations and Haitian communities, findings from the Jackson Heart Study similarly reported associations between DS, socioeconomic adversity, lower physical activity, and greater comorbidity burden.^44^ Our study complements this literature, even in the absence of traditional risk factors like smoking, obesity, and hypertension, individuals with moderate to severe DS are more likely to experience CVD. These findings underscore the need to address broader SDoH, as outlined in the ecological model,^1^ to mitigate CVD risk in populations with depression, even in the absence of traditional risk factors.

We also explored whether the relationship between DS and CVD differed by sex. In our study, females consistently exhibited higher prevalence of moderate to severe DS than males (Fig. S3), and a stronger association between DS and CVD was observed among women. Although neither the multiplicative interaction term nor the additive-scale RERI reached statistical significance, associations between DS and CVD were consistently stronger among women across strata.

Across different settings, women consistently exhibit higher rates of depression than men, making them more vulnerable to cardiovascular diseases.^4,8^ Women also experience nearly twice the incidence of CVD-related deaths compared to men.^4,45^ However, the evidence on sex differences in the DS–CVD relationship remains mixed. Our findings are consistent with some studies but contrast with others. Two prospective cohort studies from Asia also reported stronger associations between DS and incident CVD among women. Although these studies were conducted in Japan (high-income country) and China (upper-middle-income country), the study from Guizhou Province in Southwest China was conducted in one of the country’s least economically developed regions, where limited healthcare access and socioeconomic adversity may share contextual similarities with Haiti.^10,11^ In contrast, studies such as REGARDS and the Jackson Heart Study reported stronger effects in men. A meta-analysis of 17 prospective studies found no significant sex differences, highlighting the complexity of this relationship.^11^ These discrepancies may reflect differences in study populations, study designs, or unmeasured confounding. Contextual factors, such as healthcare access, cultural norms, and economic adversity in Haiti compared to high-income settings, may also have shaped the sex-specific effect. Additionally, women with depression are more likely to experience traditional cardiovascular risk factors like hypertension, diabetes, and obesity, and tend to report more persistent and severe depression related symptoms than men.^10,46–48^ Socioeconomic inequalities, including disparities in education and household income further compound these risks.^47^

Although the mechanisms underlying these potential differences are not understood, they likely involve biological factors (hormonal fluctuations, inflammatory pathways) and socioeconomic disparities including unequal access to healthcare and disproportionate caregiving burdens. Depression prevalence increases markedly in women after mid-puberty, likely due to a combination of hormonal changes and psychosocial stressors, while no differences are observed between boys and girls during childhood and early puberty.^4,9^

We hypothesized a stronger association between DS and CVD among females, and although the interaction term on the multiplicative scale was not statistically significant, associations between DS and CVD were consistently stronger among women, although the EMM on the additive scale did not reach statistical significance. Furthermore, our findings highlight the need for integrated public health strategies that address DS as a modifiable CVD risk factor, by prioritizing mental health screening, equitable access to care, and CVD prevention efforts, particularly among women in resource-limited settings.

This study has several notable strengths. First, it used the PHQ-9, a validated instrument translated into Haitian Creole. Second, it integrated SDoH into the analysis to provide a comprehensive understanding of the broader context linking depression and CVD. Finally, it assessed sex-specific effects on both additive and multiplicative scales enhancing methodological rigor and interpretation.

However, this study also has limitations. Its cross-sectional design precludes establishing causality. The generalizability of findings may be limited to urban settings and may not extend to rural areas of Haiti. Although PHQ-9 is validated, it remains a self-reported screening tool and was administered by community health and social workers rather than self-administered, potentially introducing bias. Furthermore, unmeasured confounding such as healthcare access, health literacy, family history of depression or CVD, or exposure to neighborhood violence may have influenced the observed associations. Although some covariate data were missing, results from multiple imputation analyses were consistent with the complete-case findings, suggesting minimal influence of missing data on the observed associations. (Supporting information, Table S2) Finally, individuals with more severe DS may have been less likely to enroll, introducing potential selection bias.

To address the dual burden of mental illness and CVD in Haiti, integrated public health strategies are urgently needed. These should prioritize mental health screening in primary care, address upstream drivers such as poverty and chronic stress, and ensure equitable access to care.

## Conclusion

This study highlights a strong association between moderate to severe DS and increased CVD risk in a high-risk urban population in Haiti, particularly among females. These findings reinforce the importance of depression as a modifiable CVD risk factor, emphasizing the need for routine depression screening in primary care alongside standard CVD assessments.

Given the interplay between depression, social determinants of health, and cardiovascular risk, integrating mental health education, awareness campaigns, and accessible healthcare services into public health strategies is crucial. Primary care physicians should prioritize CVD prevention in individuals with DS, even in the absence of traditional cardiovascular risk factors. Future research should address study limitations and employ longitudinal approaches to better understand the causal pathways linking DS and CVD.

## Data Availability

De-identified individual participant data underlying the findings reported in this manuscript have been deposited in Digital Commons Data@Becker and made available through controlled access under a Data Use Agreement (https://doi.org/10.17632/5bmc66w395). To protect participant confidentiality, only aggregated summary data will be shared openly.

## Supplemental Tables

**Table S1.**
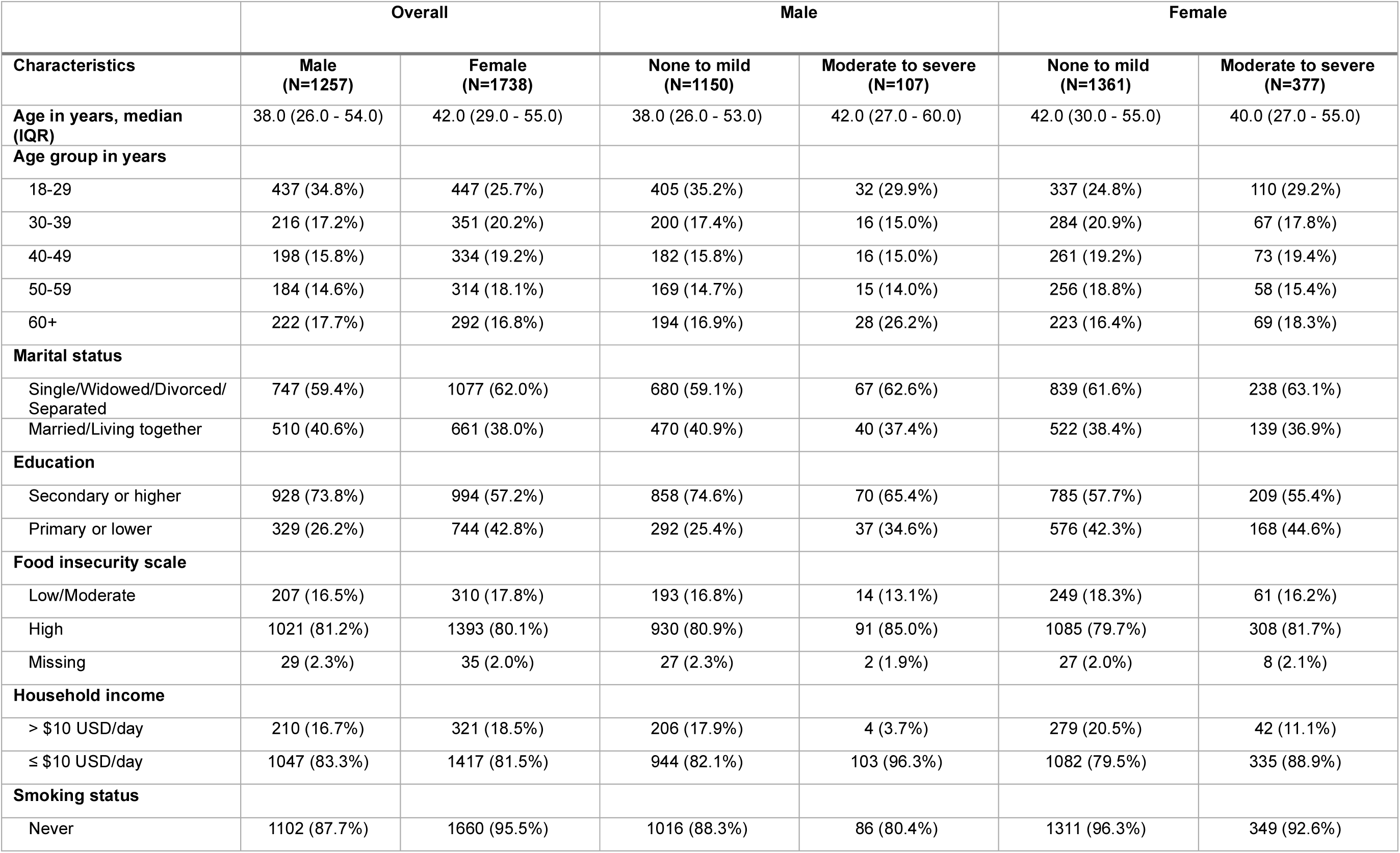
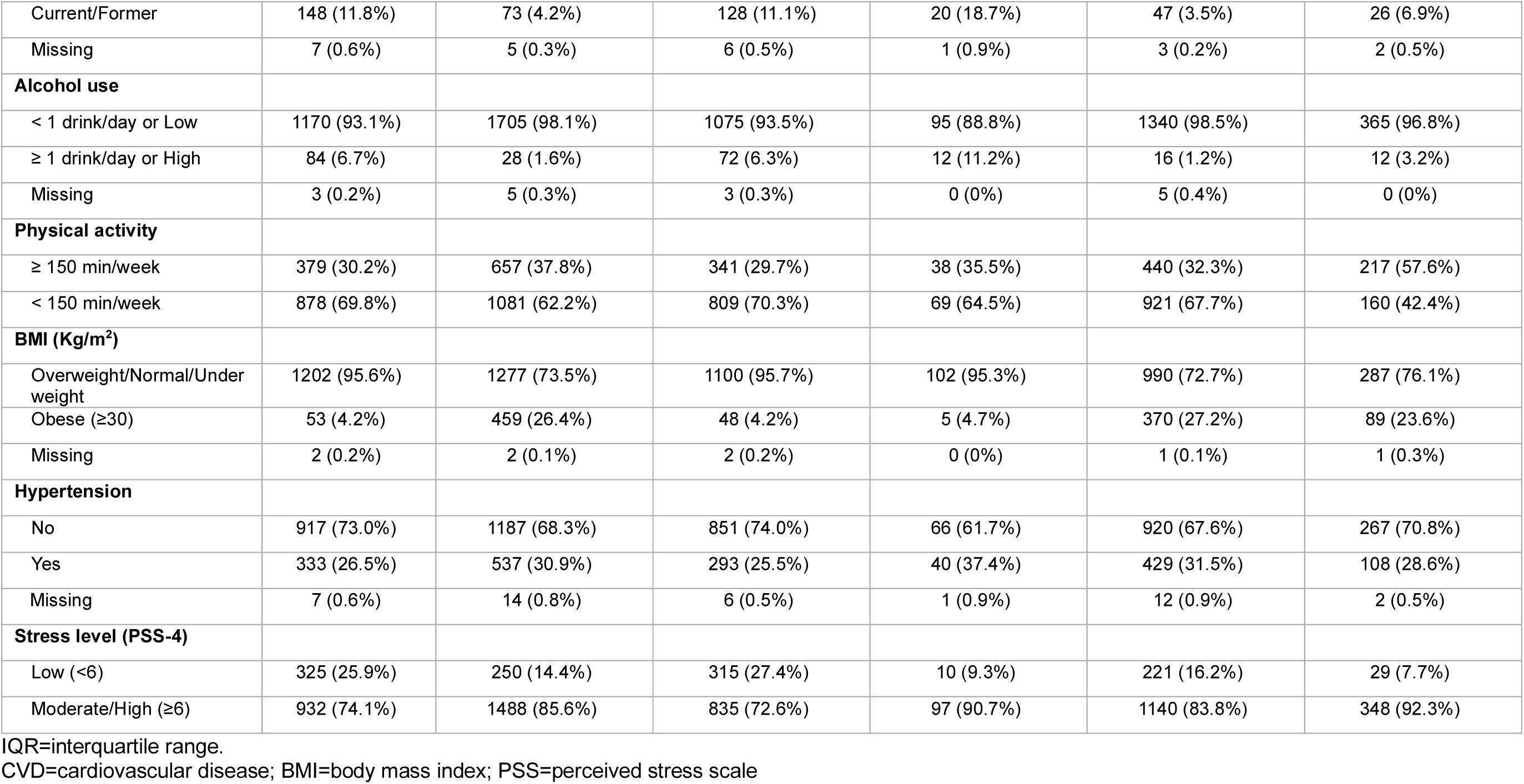
Participant Characteristics by Depressive Symptoms (DS) Level and Sex in the HCVD Cohort (n=2995), 2019-2021, Port-au-Prince, Haiti.

**Table S2.**
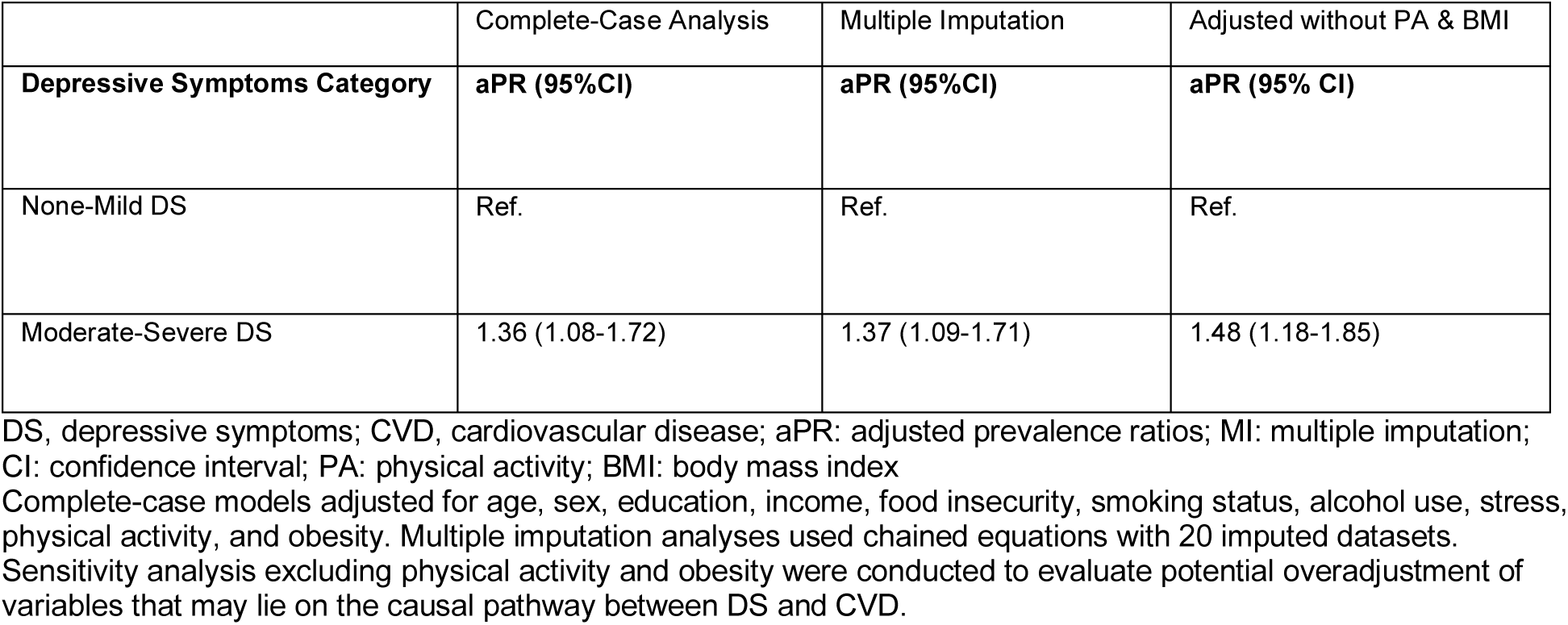
Sensitivity Analysis of the Association between Depressive Symptoms (DS) and Prevalent CVD using Complete-case Analysis, Multiple Imputation and Alternative Covariate Adjustment Strategies in the HCVD Cohort, 2019–2021, Port-au-Prince, Haiti.

---

**Fig. 1.**
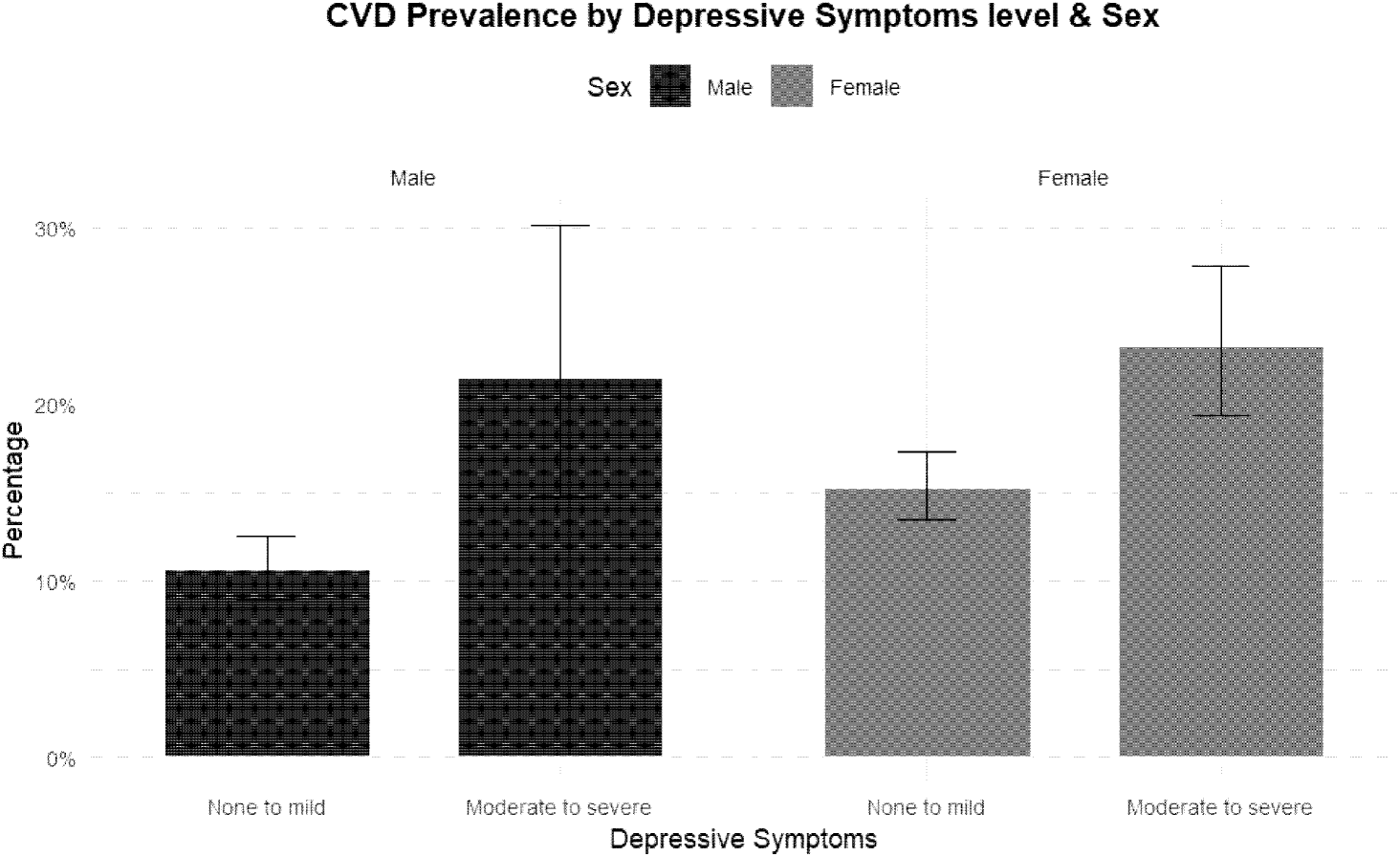
Cardiovascular Disease (CVD) Prevalence by Depressive Symptoms (DS) level and sex in HCVD Cohort, March 2019–August 21.

**Fig. S1.**
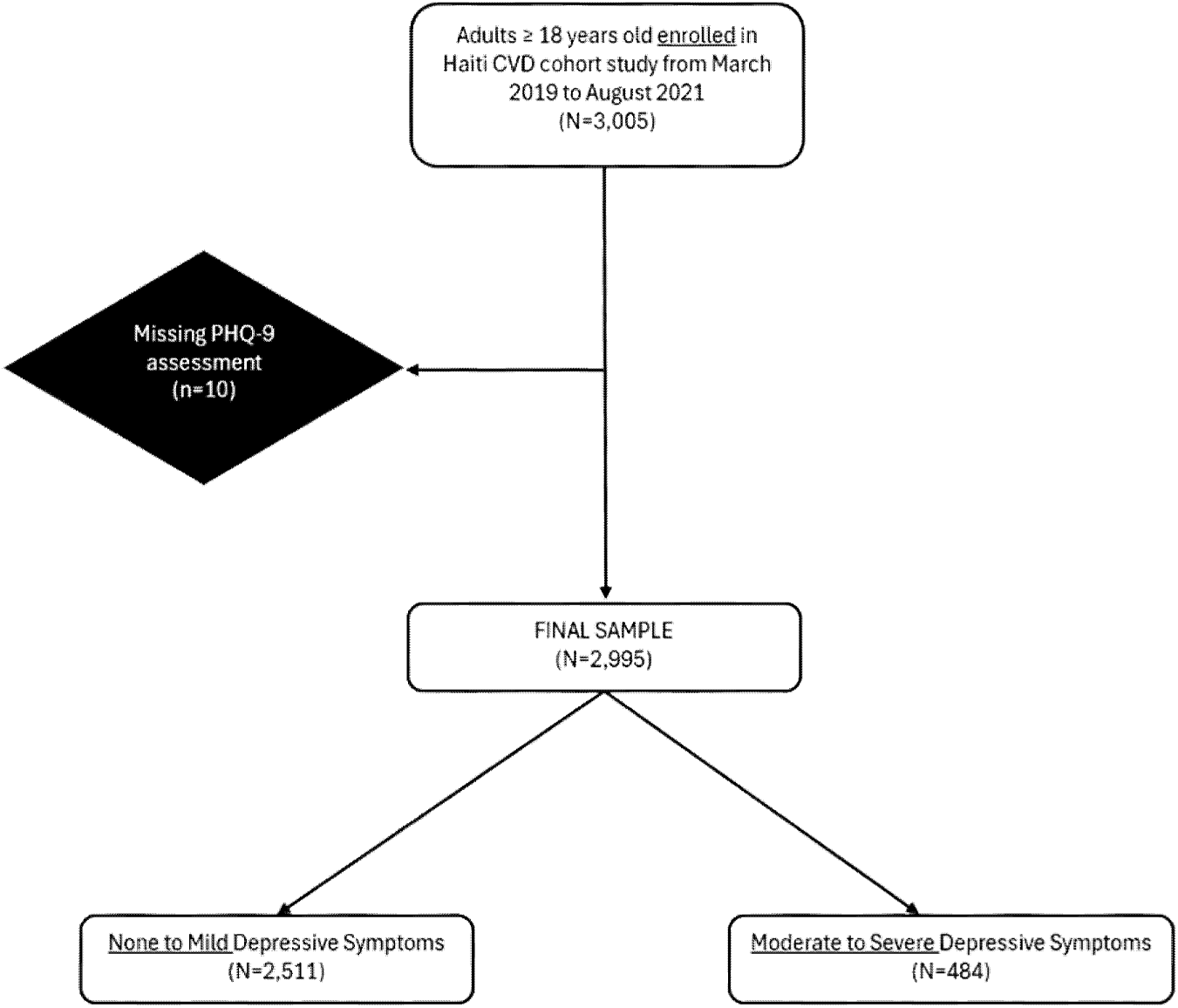
Study diagram flow: Participants Inclusion and Exclusion

**Fig. S2.**
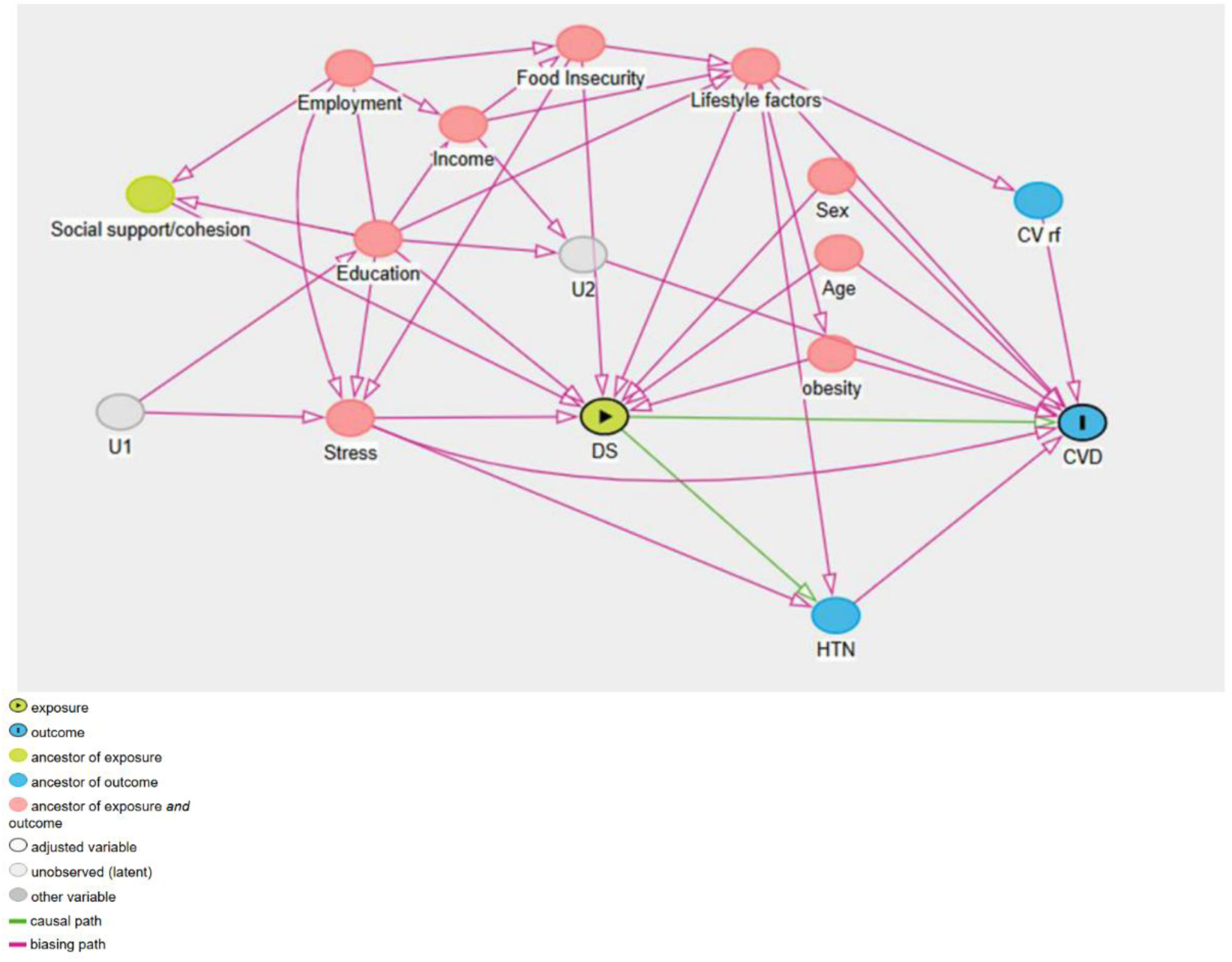
Directed-acyclic graph (DAG) Showing the Relationship Between DS and Prevalent CVD in Adults

**Fig. S3.**
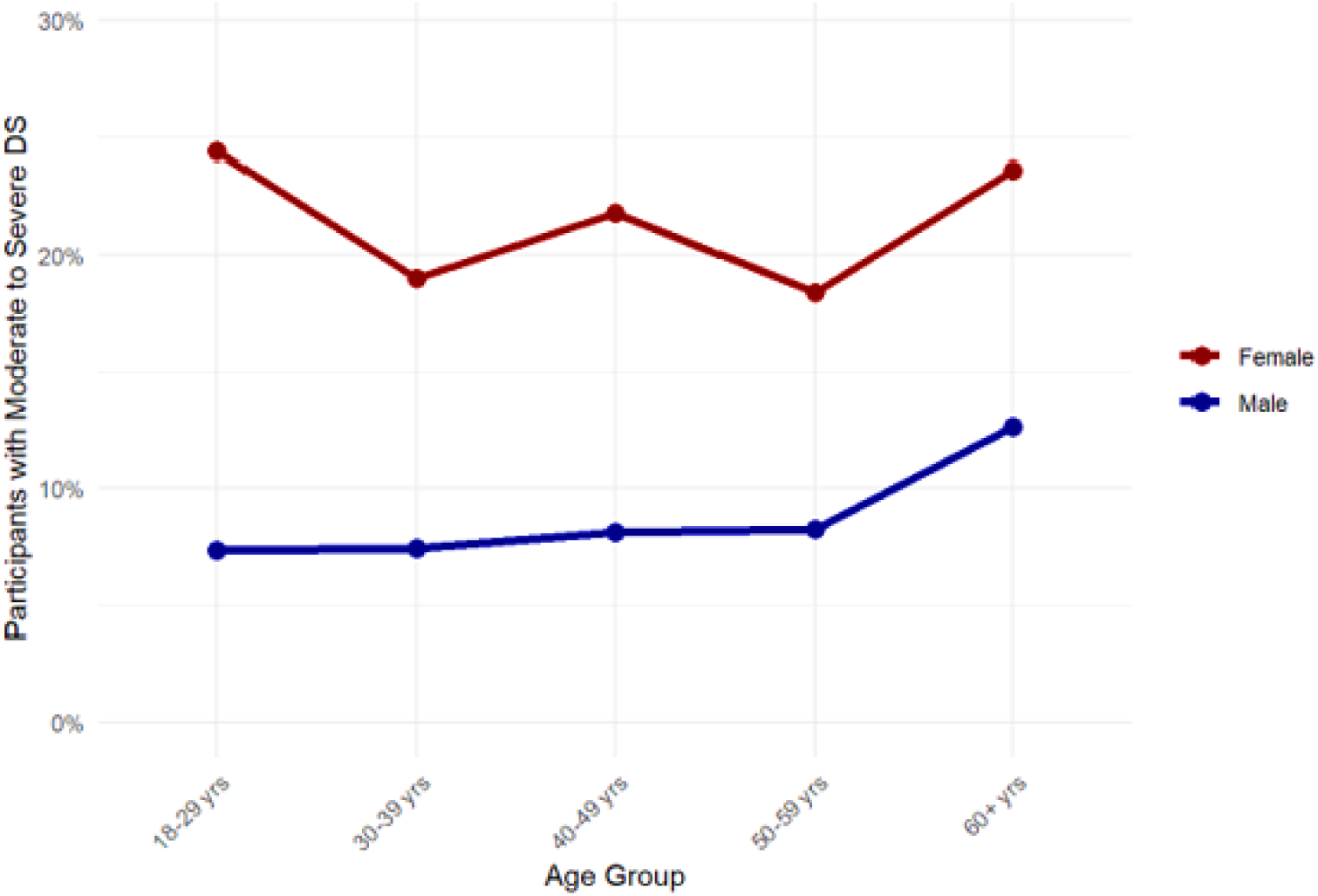
Prevalence of Moderate to Severe Depressive Symptoms (DS) by Age Group and Sex in HCVD Cohort, Haiti

