## Supporting Information for "Depressive symptoms and Prevalent Cardiovascular Disease: A Cross-Sectional Analysis in Port-au-Prince, Haiti"

**Authors and Affiliations**

Daniella Myriam Pierre*, MD, MPH, CPH^1,2^

Rehana Rasul, MA, MPH^1,2^

Reichling St. Sauveur, MD^3^

Kelly Celestin, MD^3^

Vanessa Rouzier, MD^3,4^

Erline Hilaire^3^

Marie Marcelle Deschamps, MD^3^

Jean William Pape, MD^3,4^

Lily D. Yan, MD, MSc^4^

Anju Ogyu^4^

Catherine Bennett^4^

Margaret L. McNairy, MSc, MD^4,5^

Rodney Sufra, MD^3^

Denis Nash, PhD, MPH^1,2^

* first author

1. Department of Epidemiology and Biostatistics, Graduate School of Public Health and Health Policy, City University of New York, New York, NY 10027, USA

2. Institute of Implementation Science in Population Health, City University of New York, New York, NY 10027, USA

3. Haitian Group for the Study of Kaposi’s Sarcoma and Opportunistic Infections (GHESKIO), 33 Boulevard Harry Truman, Port-au-Prince 6110, Haiti

4. Center for Global Health, Weill Cornell Medicine, 402 East 67th Street, New York, NY 10065, USA

5. Division of General Internal Medicine, Department of Medicine, Weill Cornell Medicine, 525 East 68th Street, Box 331, New York, NY, 10065, USA

**Corresponding Author**

Daniella Myriam Pierre, MD, MPH, CPH

Department of Epidemiology and Biostatistics, Graduate School of Public Health and Health Policy, City University of New York, 55 West 125th Street, New York, NY 10027, US

**Table of Contents**

1- Supplementary Methods

- Adjudication appendix……………………………………………………………………… 3
  - Appendix S1 Table 1: Adjudication criteria for prevalent CVD Events in the Haiti Cardiovascular Disease Cohort
  - Appendix S1 Table 2: Prevalence of adjudicated cardiovascular disease components in the Haiti Cardiovascular Disease Cohort (previously reported as Prevalence of CVD by sex and age^1^

2- Statistical Appendix…………………………………………………………………………...... 9
3- Supplemental tables………………………………………………………………………….. 11

- Table S1: Participant Characteristics by Depressive Symptoms (DS) Level and Sex in the HCVD Cohort (n=2995), 2019-2021, Port-au-Prince, Haiti
- Table S2: Sensitivity Analysis of the Association between Depressive Symptoms (DS) and Prevalent CVD using Complete-case Analysis, Multiple Imputation and Alternative Covariate Adjustment Strategies in the HCVD Cohort, 2019–2021, Port-au-Prince, Haiti

4- Supplemental figures………………………………………………………………………….14

- Fig S1: Study diagram flow, Participants inclusion and exclusion
- Fig S2: Directed-acyclic graph (DAG) Showing the Relationship between Depressive Symptoms (DS) and Prevalent CVD in Adults
- Fig S3: Prevalence of Moderate to Severe Depressive Symptoms (DS) by Age Group and Sex in HCVD Cohort, Haiti

**CVD Adjudication**

The adjudication procedures used in this analysis were previously developed and reported for the Haiti Cardiovascular Disease Cohort. The following summary is reproduced and adapted from the published adjudication appendix to facilitate reproducibility and address reviewer requests for detailed outcome definitions. Potential cases were identified using participant-reported symptoms, medical history, physical examination findings, imaging studies, and laboratory data. All potential cases were reviewed by a three-physician adjudication committee and classified as definite, probable, or possible cardiovascular disease events. Only definite and probable events were included in the present analysis.^1^

**Appendix S1 Table 1.-** **Adjudication criteria for prevalent CVD Events in the Haiti Cardiovascular Disease Cohort**

*Adapted from [Spectrum of prevalent cardiovascular diseases in urban Port-au-Prince, Haiti]^1^. Reproduced with permission where applicable.*

| **CVD Event** | **Category** | **Adjudication Criteria** |
| --- | --- | --- |
| Heart Failure | Definite | 1. Clinical signs and symptoms compatible with left or right sided heart failure (e.g., dyspnea, rales, S3, decreased exercise tolerance, volume overload such a peripheral edema, worsened end-organ perfusion), without an alternative explanation, with **at least one of the following**: ^2,3^   a. Radiographic evidence including hemodynamic measurements, radionucleotide ventriculography, echocardiogram, cardiac catheterization, or multiple gated acquisition scan showing: i. for Heart Failure with reduced Ejection Fraction (HFrEF) a decreased ejection fraction of less than or equal to 40% OR ii. for Heart Failure with mildly reduced Ejection Fraction (HFmrEF) a decreased ejection fraction of 41-49% OR iii. for Heart Failure with preserved Ejection Fraction (HFpEF) an ejection fraction of greater or equal than 50%, AND objective evidence of cardiac structural and/or functional abnormalities consistent with the presence of LV diastolic dysfunction / raised LV filing pressures (any of: LV mass index ≥95 g/m^2^ females or ≥115 g/m^2^ males, E/e’ ratio at rest > 9, PA systolic pressure > 35 mmHg, TR velocity at rest > 2.8 m/s, Mitral E > 90, chest X-ray showing cardiomegaly, EKG with LV hypertrophy or atrial fibrillation) b. Laboratory evidence including elevated BNP level or pro-BNP OR   c. Documentation of treatment for HF |
| Heart Failure | Probable | 1. **At least one of the following**:   a. Clinical signs and symptoms compatible with left or right sided heart failure (e.g., dyspnea, rales, S3, decreased exercise tolerance, volume overload such a peripheral edema, worsened end-organ perfusion), without an alternative explanation, but with no available data from radiographic, laboratory, or treatment OR  b. Patient self-report of physician telling them that they have a diagnosis of heart failure but medical records are not available      AND  2. No alternative diagnosis as cause of symptoms  AND  3. Physician diagnosis of heart failure |

| Stroke | Definite | **All of the following** 2,4,5   1. **Either:**    a. Rapidly developing clinical signs of focal (or global) disturbance of cerebral function, with symptoms lasting 24 hours or longer or leading to death, with no apparent cause other than of vascular origin OR    b. Death certificate or death NOTE from medical record listing stroke as cause of death   AND  2. Physical exam findings consistent with focal or global disturbance of cerebral function with no apparent cause other than of vascular origin  AND  3. Demonstrable lesion compatible with acute ischemic or hemorrhagic stroke on a CT or MRI   i. acute ischemic stroke   1. Focal brain deficit without CT or LP evidence of blood, except mottled cerebral pattern. Either decreased density by CT in a compatible location or a negative CT or none done. OR  2.  Surgical evidence of ischemic infarction   ii. acute hemorrhagic stroke   1. Blood in subarachnoid space or intraparenchymal hemorrhage by CT. (Intraparenchymal blood must be dense, not mottled-mixed hyper- and hypodensity.) OR  2. Bloody spinal fluid by lumbar puncture (bloody CSF means > 100 cells/mm3. The LP is thought to be a non-traumatic and counts in the last tube are similar to those in the first tube (no clearing) or xanthochromia when the specimen is spun down.) OR  3. Surgical evidence of hemorrhage as cause of clinical syndrome |
| --- | --- | --- |
| Stroke | Probable | **All of the following** 2,4,5   1. **Either:** a. Rapidly developing clinical signs of focal (or global) disturbance of cerebral function, with symptoms lasting 24 hours or longer or leading to death, with no apparent cause other than of vascular origin OR b. Death certificate or death NOTE from medical record listing stroke as cause of death   AND   2. Physical exam findings consistent with focal or global disturbance of cerebral function with no apparent cause other than of vascular origin   AND   3. Lack of data from imaging test like CT head or MRI   OR   4. Patient reported past medical history |
| Transient Ischemic Attack |  | **1. At least one of the following: ^2,4^**  a. Rapidly developing clinical signs of focal (or global) disturbance of cerebral function, with symptoms lasting < 24 hours with no apparent cause other than of vascular origin OR  b. Patient self-report of physician telling them they have a diagnosis of TIA but medical records not available |
| Prior Myocardial Infarction | Definite | **1. At baseline, at least one of the following ^3^**  a. ECG indicating findings consistent with old myocardial infarction. For example, Q wave present in 2 or more contiguous leads and with either duration greater than or equal to 40 msec or amplitude greater than ¼ R wave. OR  b. Imaging evidence of loss of viable myocardium or regional wall motion abnormality |
| Prior Myocardial Infarction | Probable | **1. Patient self-report of prior MI diagnosis** |
| Angina | Definite ^5^ | Both of the following:  1. History of chest discomfort associated with exertion or excitement and alleviated with rest. May be described as pain but more frequently as heaviness, pressure, squeezing, or choking sensation. May radiate to the left shoulder, down the arm, back, neck, or jaw. Rose Angina questionnaire ((B1 or B4)+B2 OR (B1 or B4)+B3).  AND   2. At **least one of the following**:  a. ECG consistent with ischemia (T wave inversions, ST depressions or elevations, pathologic Q waves) OR b. Stress test finding consistent with ischemia OR c. Angiogram or other imaging test of coronary arteries demonstrating significant occlusion, and other etiologies of the presenting signs and symptoms unlikely. |
| Angina | Probable ^5^ | All of the following:  1. History of chest discomfort associated with exertion or excitement and alleviated with rest. May be described as pain but more frequently as heaviness, pressure, squeezing, or choking sensation. May radiate to the left shoulder, down the arm, back, neck, or jaw. Rose Angina questionnaire ((B1 or B4)+B2 OR (B1 or B4)+B3).  AND  2. Lack of data from ECG, stress test, angiogram, or other imaging test demonstrating ischemia or coronary artery disease  AND  3. Physician diagnosis of angina without another cause of chest pain  OR  1. Patient reported past medical history |

**Appendix S1 Table 2.-** **Component-specific frequencies of adjudicated cardiovascular disease outcomes in the Haiti Cardiovascular Disease Cohort**

| **Component** | **N (%)** |
| --- | --- |
| Heart failure | 352 (11.7%) |
| Stroke | 77 (2.6%) |
| Transient Ischemic Attack (TIA) | 14 (0.5%) |
| Myocardial Infarction (MI) | 30 (1.0%) |
| Angina | 65 (2.2%) |
| Any CVD (primary outcome) | 442 (14.7%) |
| CVD (-Angina/TIA) | 410 (13.7%) |

*These prevalence estimates were previously reported in the parent Haiti Cardiovascular Disease Cohort publication and are presented here to provide component-specific frequencies underlying the composite CVD outcome used in the present analysis. Participants could contribute to more than one CVD category; therefore, component frequencies do not sum to the total number of participants with CVD.^1^*

**-**

**Statistical Appendix**

**Model Specification for Prevalence Ratios**

To estimate the prevalence ratio (PR) of CVD associated with depressive symptoms, we used Poisson regression models with a log link, both with and without an interaction term for sex. This was done for both crude and adjusted models.

1- For the crude models, the following model specification was used first:

*log(CVDpr_i_) = β_0_ + β_1_DS_i_ for i=1,…,n of n…*

Secondly, adjusted only for age to account for potential confounding by age, then an interaction term (DS * sex) was introduced to evaluate whether the association between DS and CVD differed by sex in the crude model (Table 2, Table 3).

2- For the adjusted model, a multivariable Poisson regression model was then used to estimate the adjusted prevalence ratio (aPR), adjusting for age, sex, income, food insecurity, alcohol consumption, smoking, stress, physical activity, and obesity (Table 4). The model specification equation was:

*log(CVDpr_i_) = β_0_ + β_1_DS_i_ + β_2_sex_i_ + β_3_age_i_ + β_4_education_i_ + β_5_income_i_ + β_6_FI_i_ + β_7_alcohol_i_ + β_8_smoking_i_ + β_9_pactivity_i_ + β_10_stress_i_ + β_11_obesity_i_*

**Effect Modification on the Multiplicative Scale**

To assess whether sex modifies the relationship between depressive symptoms and CVD on the multiplicative scale, an interaction term (DS * sex) was added to the adjusted regression model:

*log(CVDpr_i_) = β_0_ + β_1_DS_i_ + β_2_sex_i_ + β_3_DS_i_∗sex_i_ + β_4_age_i_ + β_5_education_i_ + β_6_income_i_ + β_7_FI_i_ + β_8_alcohol_i_ + β_9_smoking_i_ + β_10_pactivity_i_ + β_11_stress_i_ + β_12_obesity_i_*

A statistically significant p-value for the interaction term (β_3_) was considered as evidence of effect modification by sex on the multiplicative scale. To further investigate these sex-specific effects, stratified analyses were performed, and estimated marginal means (EMMs) were computed(Table 4).

**Effect Modification on the Additive Scale**

On the additive scale, EMM was measured with Rothman’s formula for Relative Excess Risk due to Interaction (RERI), and the 95% CI was calculated with the Epinet spreadsheet for the RERI:

𝑅𝐸𝑅𝐼_𝑅𝑅_ = 𝑅𝑅_11_ − 𝑅𝑅_10_ − 𝑅𝑅_01_ + 1

Since prevalence ratios (PRs) were used in this study design instead of risk ratios (RRs), the formula was adapted accordingly:

𝑅𝐸𝑅𝐼_𝑃𝑅_ = 𝑃𝑅_11_ − 𝑃𝑅_10_ − 𝑃𝑅_01_ + 1

Where:

𝑃𝑅_11_ = Prevalence ratio for participants with moderate to severe DS and female,

𝑃𝑅_10_ = Prevalence ratio for participants with moderate to severe DS and male,

𝑃𝑅_01_ = Prevalence ratio for participants with none to mild DS and female,

𝑃𝑅_00_ = Reference group, typically set to 1

**Interpreting RERI**

RERI = 0 → No interaction (perfect additivity).

RERI > 0 → Positive interaction (more than additivity), meaning the combined effect is greater than expected under additivity.

RERI < 0 → Negative interaction (less than additivity), meaning the combined effect is lower than expected under additivity.

**Supplemental Tables**

**Table S1.-** Participant Characteristics by Depressive Symptoms (DS) Level and Sex in the HCVD Cohort (n=2995), 2019-2021, Port-au-Prince, Haiti

|  | Overall | | Male | | Female | |
| --- | --- | --- | --- | --- | --- | --- |
| Characteristics | **Male**  **(N=1257)** | **Female**  **(N=1738)** | **None to mild**  **(N=1150)** | **Moderate to severe**  **(N=107)** | **None to mild**  **(N=1361)** | **Moderate to severe**  **(N=377)** |
| Age in years, median (IQR) | 38.0 (26.0 - 54.0) | 42.0 (29.0 - 55.0) | 38.0 (26.0 - 53.0) | 42.0 (27.0 - 60.0) | 42.0 (30.0 - 55.0) | 40.0 (27.0 - 55.0) |
| Age group in years |  |  |  |  |  |  |
| 18-29 | 437 (34.8%) | 447 (25.7%) | 405 (35.2%) | 32 (29.9%) | 337 (24.8%) | 110 (29.2%) |
| 30-39 | 216 (17.2%) | 351 (20.2%) | 200 (17.4%) | 16 (15.0%) | 284 (20.9%) | 67 (17.8%) |
| 40-49 | 198 (15.8%) | 334 (19.2%) | 182 (15.8%) | 16 (15.0%) | 261 (19.2%) | 73 (19.4%) |
| 50-59 | 184 (14.6%) | 314 (18.1%) | 169 (14.7%) | 15 (14.0%) | 256 (18.8%) | 58 (15.4%) |
| 60+ | 222 (17.7%) | 292 (16.8%) | 194 (16.9%) | 28 (26.2%) | 223 (16.4%) | 69 (18.3%) |
| Marital status |  |  |  |  |  |  |
| Single/Widowed/Divorced/Separated | 747 (59.4%) | 1077 (62.0%) | 680 (59.1%) | 67 (62.6%) | 839 (61.6%) | 238 (63.1%) |
| Married/Living together | 510 (40.6%) | 661 (38.0%) | 470 (40.9%) | 40 (37.4%) | 522 (38.4%) | 139 (36.9%) |
| Education |  |  |  |  |  |  |
| Secondary or higher | 928 (73.8%) | 994 (57.2%) | 858 (74.6%) | 70 (65.4%) | 785 (57.7%) | 209 (55.4%) |
| Primary or lower | 329 (26.2%) | 744 (42.8%) | 292 (25.4%) | 37 (34.6%) | 576 (42.3%) | 168 (44.6%) |
| Food insecurity scale |  |  |  |  |  |  |
| Low/Moderate | 207 (16.5%) | 310 (17.8%) | 193 (16.8%) | 14 (13.1%) | 249 (18.3%) | 61 (16.2%) |
| High | 1021 (81.2%) | 1393 (80.1%) | 930 (80.9%) | 91 (85.0%) | 1085 (79.7%) | 308 (81.7%) |
| Missing | 29 (2.3%) | 35 (2.0%) | 27 (2.3%) | 2 (1.9%) | 27 (2.0%) | 8 (2.1%) |
| Household income |  |  |  |  |  |  |
| > $10 USD/day | 210 (16.7%) | 321 (18.5%) | 206 (17.9%) | 4 (3.7%) | 279 (20.5%) | 42 (11.1%) |
| ≤ $10 USD/day | 1047 (83.3%) | 1417 (81.5%) | 944 (82.1%) | 103 (96.3%) | 1082 (79.5%) | 335 (88.9%) |
| Smoking status |  |  |  |  |  |  |
| Never | 1102 (87.7%) | 1660 (95.5%) | 1016 (88.3%) | 86 (80.4%) | 1311 (96.3%) | 349 (92.6%) |
| Current/Former | 148 (11.8%) | 73 (4.2%) | 128 (11.1%) | 20 (18.7%) | 47 (3.5%) | 26 (6.9%) |
| Missing | 7 (0.6%) | 5 (0.3%) | 6 (0.5%) | 1 (0.9%) | 3 (0.2%) | 2 (0.5%) |
| Alcohol use |  |  |  |  |  |  |
| < 1 drink/day or Low | 1170 (93.1%) | 1705 (98.1%) | 1075 (93.5%) | 95 (88.8%) | 1340 (98.5%) | 365 (96.8%) |
| ≥ 1 drink/day or High | 84 (6.7%) | 28 (1.6%) | 72 (6.3%) | 12 (11.2%) | 16 (1.2%) | 12 (3.2%) |
| Missing | 3 (0.2%) | 5 (0.3%) | 3 (0.3%) | 0 (0%) | 5 (0.4%) | 0 (0%) |
| Physical activity |  |  |  |  |  |  |
| ≥ 150 min/week | 379 (30.2%) | 657 (37.8%) | 341 (29.7%) | 38 (35.5%) | 440 (32.3%) | 217 (57.6%) |
| < 150 min/week | 878 (69.8%) | 1081 (62.2%) | 809 (70.3%) | 69 (64.5%) | 921 (67.7%) | 160 (42.4%) |
| BMI (Kg/m^2^) |  |  |  |  |  |  |
| Overweight/Normal/Underweight | 1202 (95.6%) | 1277 (73.5%) | 1100 (95.7%) | 102 (95.3%) | 990 (72.7%) | 287 (76.1%) |
| Obese (≥30) | 53 (4.2%) | 459 (26.4%) | 48 (4.2%) | 5 (4.7%) | 370 (27.2%) | 89 (23.6%) |
| Missing | 2 (0.2%) | 2 (0.1%) | 2 (0.2%) | 0 (0%) | 1 (0.1%) | 1 (0.3%) |
| Hypertension |  |  |  |  |  |  |
| No | 917 (73.0%) | 1187 (68.3%) | 851 (74.0%) | 66 (61.7%) | 920 (67.6%) | 267 (70.8%) |
| Yes | 333 (26.5%) | 537 (30.9%) | 293 (25.5%) | 40 (37.4%) | 429 (31.5%) | 108 (28.6%) |
| Missing | 7 (0.6%) | 14 (0.8%) | 6 (0.5%) | 1 (0.9%) | 12 (0.9%) | 2 (0.5%) |
| Stress level (PSS-4) |  |  |  |  |  |  |
| Low (<6) | 325 (25.9%) | 250 (14.4%) | 315 (27.4%) | 10 (9.3%) | 221 (16.2%) | 29 (7.7%) |
| Moderate/High (≥6) | 932 (74.1%) | 1488 (85.6%) | 835 (72.6%) | 97 (90.7%) | 1140 (83.8%) | 348 (92.3%) |

IQR=interquartile range.

CVD=cardiovascular disease; BMI=body mass index; PSS=perceived stress scale

**Table S2.-** Sensitivity Analysis of the Association between Depressive Symptoms (DS) and Prevalent CVD using Complete-case Analysis, Multiple Imputation and Alternative Covariate Adjustment Strategies in the HCVD Cohort, 2019–2021, Port-au-Prince, Haiti

|  | Complete-Case Analysis | Multiple Imputation | Adjusted without PA & BMI |
| --- | --- | --- | --- |
| **Depressive Symptoms Category** | **aPR (95%CI)** | **aPR (95%CI)** | **aPR (95% CI)** |
| None-Mild DS | Ref. | Ref. | Ref. |
| Moderate-Severe DS | 1.36 (1.08-1.72) | 1.37 (1.09-1.71) | 1.48 (1.18-1.85) |

DS, depressive symptoms; CVD, cardiovascular disease; aPR: adjusted prevalence ratios; MI: multiple imputation; CI: confidence interval; PA: physical activity; BMI: body mass index

Complete-case models adjusted for age, sex, education, income, food insecurity, smoking status, alcohol use, stress, physical activity, and obesity. Multiple imputation analyses used chained equations with 20 imputed datasets. Sensitivity analysis excluding physical activity and obesity were conducted to evaluate potential overadjustment of variables that may lie on the causal pathway between DS and CVD.

**Supplemental Figures**

Fig S1: Study diagram flow, Participants inclusion and exclusion

Fig S2: Directed-acyclic graph (DAG) Showing the Relationship between Depressive Symptoms (DS) and Prevalent CVD in Adults

Fig S3: Prevalence of Moderate to Severe Depressive Symptoms (DS) by Age Group and Sex in HCVD Cohort, Haiti
